# Evaluation of the sensitivity, accuracy and currency of the Cochrane COVID-19 Study Register for supporting rapid evidence synthesis production

**DOI:** 10.1101/2021.04.21.21255874

**Authors:** Maria-Inti Metzendorf, Robin M. Featherstone

## Abstract

**Introduction:** The Cochrane COVID-19 Study Register (CCSR) is a public, continually updated, curated database of COVID-19 study references. The aim of this study-based register is to support rapid and living evidence synthesis, including a project to build an evidence ecosystem of COVID-19 research (CEOsys). In November and December 2020 we conducted an evaluation of the CCSR for CEOsys, measured its performance and identified areas for improvement.

**Methods:** For the evaluation we generated a purposive sample of 286 studies from 20 reviews to calculate the CCSR’s sensitivity (comprehensiveness), accuracy (correctly classified and linked studies) and currency (time to publish and process references).

**Results:** The CCSR had an overall sensitivity of 77.2%, with the highest sensitivity for interventional studies (94.4%) and lowest sensitivity for modelling studies (63.6%). The study register had 100% sensitivity for trial registry records, 86.5% for journal articles and 52.4% for preprints. 98.3% of references were correctly classified with regard to study type, and 93.4% with regard to study aim. 89% of studies were correctly linked. 81.4% of references were published to the register in under 30 days, with 0.5 day (median) for trial registry records, 2 days for journal articles and 56 days for preprints.

**Conclusion:** The CCSR had high sensitivity, accurate study classifications and short publishing times for journal articles and trial registry records. We identified that the CCSR’s coverage and publishing times for preprints needed improvement. Finally, the evaluation illustrated the value of a study-based register for identifying additional study references for analysis in evidence synthesis.

## 1. Introduction

In early 2020, an explosion in publication output related to the COVID-19 pandemic prompted an urgent need for an information system to support health researchers and Cochrane authors producing evidence syntheses. In March 2020 Cochrane undertook the development of the Cochrane COVID-19 Study Register (CCSR), an open-access search portal of COVID-19 primary study publicly available at covid-19.cochrane.org. On 1 April 2020 Cochrane launched the public web application of the CCSR with 868 references.^1^ Since then, daily publication of new references to the search portal has been supported by ongoing study identification and annotation. As of 8 March 2021, 48,008 studies have been published to the register.

The CCSR is a continually updated and curated collection of COVID-19 published and registered study references, including preprints. It is study-based, meaning references to the same study (e.g. press releases, trial registry records, preprints, journal pre-proofs, journal articles, retraction notices and expressions of concern) are all linked to a single study record. Study-based registers add efficiency to review production and reduce the time needed to combine different references belonging to the same study.^2,3^

The aim of the CCSR is to support rapid and living evidence synthesis, including the support of a newly created evidence ecosystem for COVID-19 research (CEOsys). CEOsys, a consortium of scientists from 21 German university medical centers and several other international partners, aims to provide researchers and guideline developers with a comprehensive, up-to-date source of curated data from clinical and public health studies. Based on the evidence assembled in the ecosystem, CEOsys is creating a series of living evidence syntheses on the most important questions on COVID-19. CEOsys evidence syntheses are subsequently translated to living recommendations to inform clinical and public health practice (publicly available at covid-evidenz.de). CEOsys focuses on the fields of diagnostics, outpatient and inpatient care, intensive and palliative care, hospital hygiene, public health and mental health.^4^

The CCSR, which serves as the primary source of information for the evidence ecosystem, was built in the Cochrane Register of Studies (CRS), a records management system and data repository, and is maintained by the Cochrane COVID-19 Study Register Centralised Search Team and associated information specialists working on the CEOsys project. The team executes daily or weekly searches, screens the results, links publications for the same studies, and classifies the references in CRS by “study reference type,” “results available,” and “study characteristics.” Automated searching progressively replaced manual searching over the course of 2020. References are first imported into a CRS segment for evaluation and classification and afterwards published to the CCSR. A complementary process of evaluating and classifying references in the CCSR segment is conducted by contributors to COVID Quest, a citizen science task hosted on Cochrane Crowd (crowd.cochrane.org).^5^ Cochrane Crowd contributors are community volunteers who assist by assessing references for eligibility in the CCSR and providing study classifications. The production process of the CCSR is depicted in Figure 1.

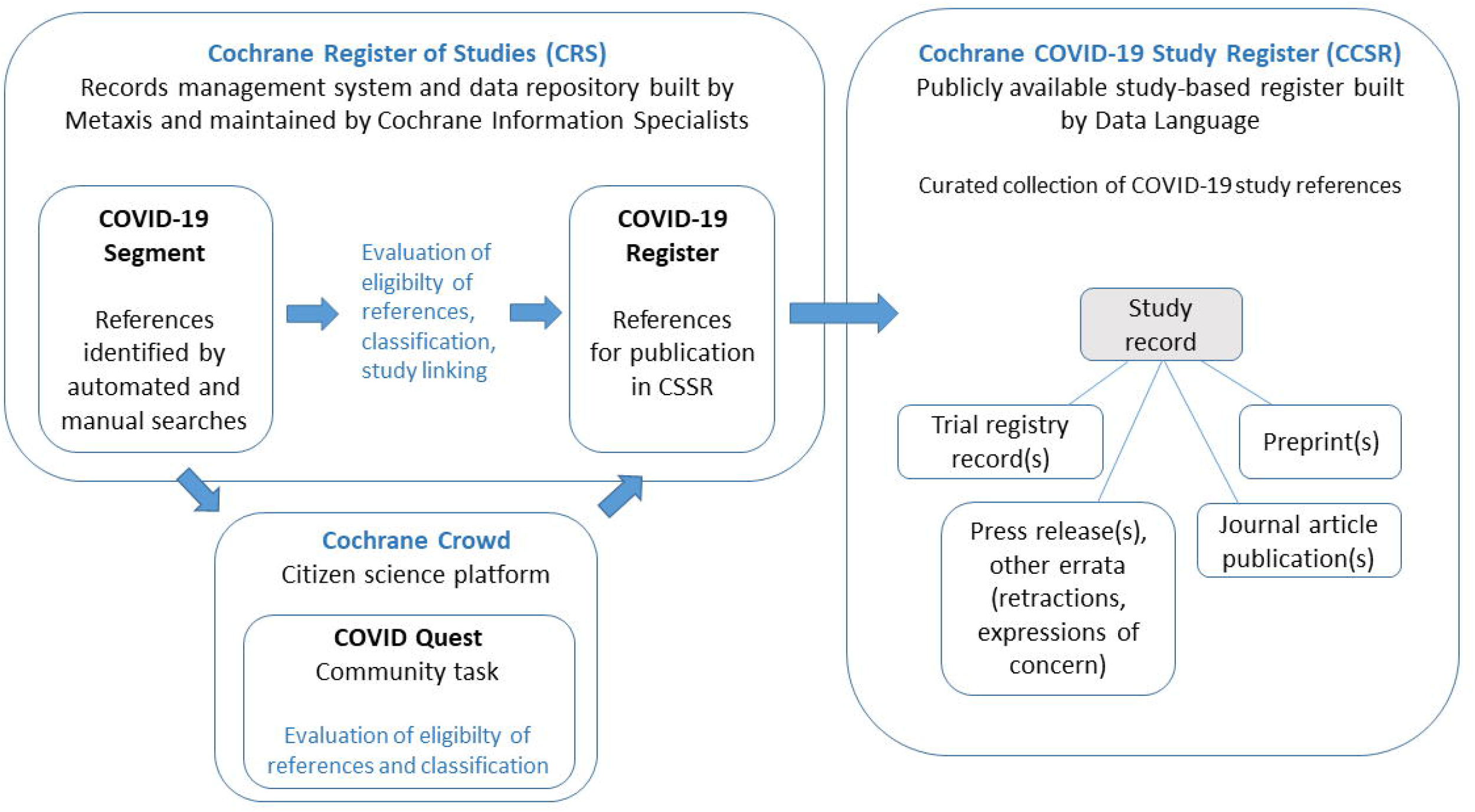

An early internal evaluation of the CCSR was conducted in mid-May 2020. It revealed the primacy of preprints (i.e., full draft research papers that are publicly shared before peer review) and letters in the growing COVID-19 evidence base. A subsequently low sensitivity (39%) of the CCSR during its first production months was found by the internal evaluation. Therefore, the Centralised Search

Team revised the CCSR’s eligibility criteria and began including both preprints and letters at the end of May 2020. In addition, the team began to capture and re-evaluate preprints and letters published between December 2019 to May 2020.

This publication describes a formal, in-depth evaluation study conducted in November and December 2020. We measured how the CCSR performed in the first half of 2020 as an information source to support COVID-19 evidence synthesis production and explored what processes should be changed to improve the study register. Our aim was to determine the sensitivity, accuracy and currency of the CCSR to support rapid and living COVID-19 review production with a focus on the CEOsys evidence ecosystem for COVID-19 research.

## 2. Methods

We prespecified our methods in a protocol published on Open Science Framework (OSF) on 20 November 2020.^6^

### 2.1. Sample

A single sample of included studies from published reviews was used to evaluate the performance of the CCSR. Given resource limitations by the investigators, we generated a purposive sample of studies from 20 COVID-19 related reviews.

Our sample aimed to include an equal number of different review types (rapid, systematic, living) and a purposive representation of 2/3 interventional (pharmacological or non-pharmacological) and 1/3 non-interventional (diagnostic, prognostic, qualitative, transmission) topics. We classified the reviews in our sample according to the six CEOsys topic areas: diagnosis, outpatient and inpatient care, intensive and palliative care, hospital hygiene, public health, or mental health. To generate our sample and ensure a balanced, unbiased set of studies we devised the following criteria.

Inclusion criteria:

- Reviews published since 1 July 2020 (for rapid and systematic reviews);
- Last update published since 1 August 2020 (for living reviews);
- Published by Cochrane or a journal with an impact factor > 4.0 according to the 2019 Journal Citation Reports category for “Medicine, General & Internal”;
- Reported a review question equivalent to one of the six CEOsys topic areas;
- Included studies on COVID-19. In the event that a review presented both direct and indirect evidence, we included only the COVID-19 related studies.

Exclusion criteria:

- Reviews published as genomic-wide association studies or prevalence studies;
- Included 0 studies or >75 studies;
- Were included in the previous internal evaluation or consulted to identify study references for the CCSR;
- Included only indirect evidence from other comparable viruses, infectious diseases or pandemics.

A PubMed search (Supplementary file 1) was used to generate reviews for the sample. It retrieved 40 records on 21 November 2020. Three additional Cochrane Reviews on COVID-19, which had not yet been indexed in PubMed, were identified via cochranelibrary.com/covid-19. Of 43 potentially relevant reviews, 31 reviews were eligible on a title/abstract basis. Eleven reviews were excluded after further inspection. Reasons:

- irrelevant topics for CEOsys = 7;
- already included in sample of previous evaluation = 1;
- 0 or more than 75 included studies = 3.

Twenty reviews were included in the final sample (Table 1).

**Table 1:**
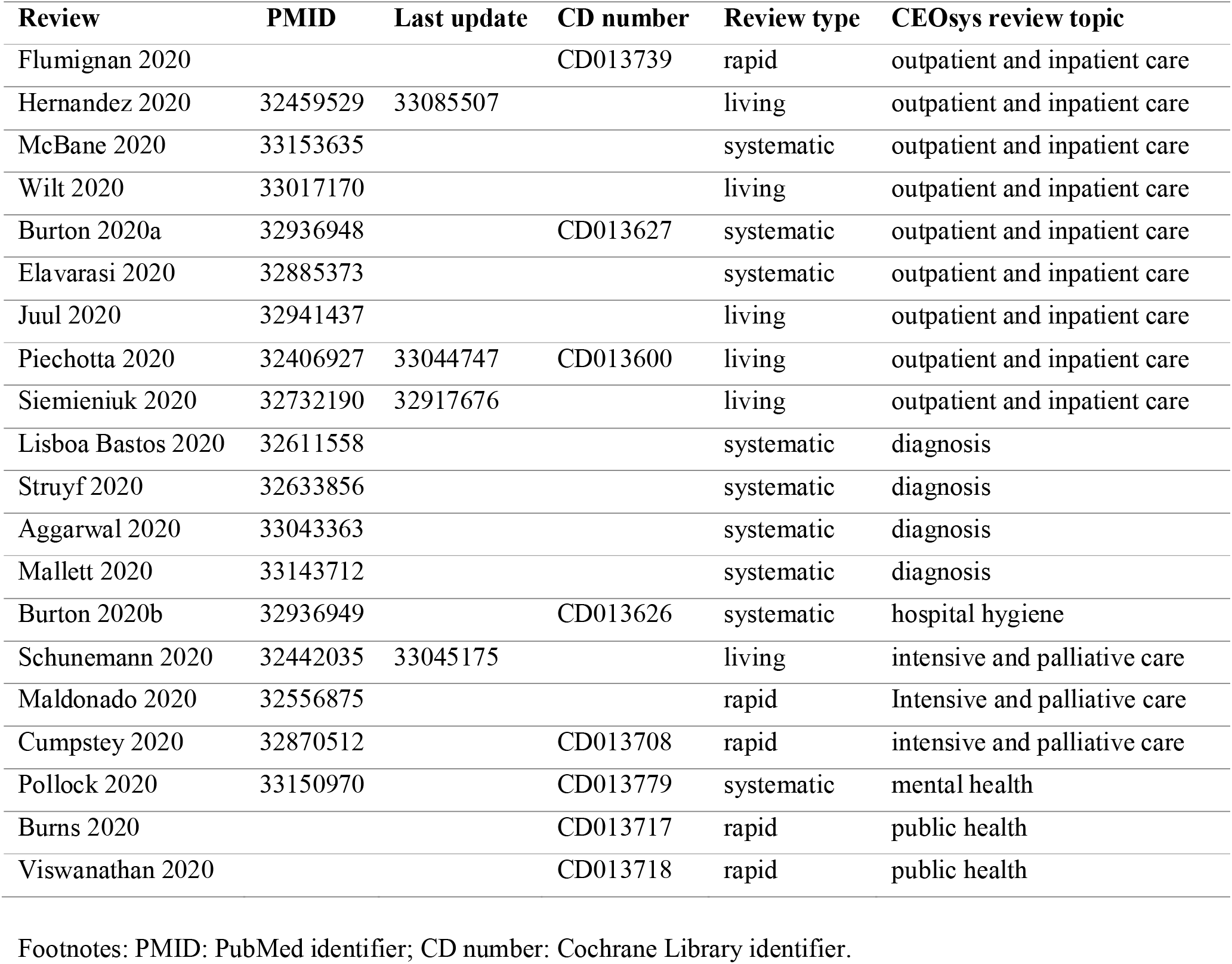
20 Reviews included in the sample.

### 2.2. Performance indicators

We used three types of performance indicators for this evaluation:

#### 1. Sensitivity (comprehensiveness of the register)

To evaluate the sensitivity of the CCSR, we used study references from included studies in the sample of reviews on COVID-19. We determined if references to these studies were contained within the CCSR. To calculate sensitivity, we did not consider the additional references included in the CCSR but based the calculation solely on those references included in the reviews. We calculated sensitivity as:

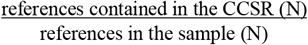

Results were calculated as a score from 0.0 to 1.0 and presented as a percentage of study references from the sample contained within the CCSR.

#### 2. Accuracy (accuracy of the classifications and linked publications)

To evaluate the accuracy of the CCSR’s classifications, we determined if references from the sample were (1) classified with the correct study type (interventional or observational), and (2) assigned a study aim that matched the aim of each included study (diagnostic/prognostic, treatment/management, health services research, etc.). We compared these classifications with details reported in the studies’ abstracts. Only if the study type and aim were not included in the abstracts did we consult the full text. We calculated the accuracy of the classifications for study type and aim as:

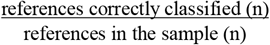

To evaluate the accuracy of linked publications (different references for the same study), we recorded if multiple references for the same study were presented in the reviews. We determined (1) if multiple references were contained in the CCSR, and (2) if they were linked via a single study reference. We also assessed if the register included linked references for the same studies that were not reported in the review. We calculated the accuracy of the linked references as:

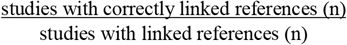

Results are presented as percentages of correctly classified study references and correctly linked references from the sample contained in the CCSR.

#### 3. Currency

In the production of the CCSR, references are first added to the segment in CRS (Figure 1). We defined “time to segment” as the time from the publication of the reference in the original source until added to the segment, reflecting the search processes. “Time from segment to register” was defined as the time it takes references to be processed and fully published to the CCSR. Both time periods, “time to segment” and “time from register to segment” constitute the publishing time. “Publishing time” is defined as the time it takes from the publication of the reference in the original source until it is fully published in the register.

To evaluate the currency of the CCSR, we recorded the time (in days), between when (1) the reference was first publicly available and when it was added to the CCSR (i.e. the publishing time), and (2) the time between when the reference was first added to the CRS’ COVID-19 segment and when it was added to the CCSR (i.e. the processing time).

### 2.3. Data collection

We independently checked each study reference from the sample to determine:

- if the reference was contained in the CCSR public search interface or the CRS segment,
- if the reference’s classifications for study type were correct,
- if the reference’s classifications for study aim were correct,
- if references for the same studies were linked correctly,
- when the references were initially available online in the original source and when they were added to the CRS segment and the CCSR.

During a primary data collection phase, a 5% sample of the references was evaluated by each investigator and reviewed by the other to ensure accuracy. All evaluation study data were recorded in Excel and are publicly available.^7^ Evaluation metrics were calculated by one investigator and verified by the other.

Data collection was carried out as planned between 21 November and 16 December 2020. 383 studies were extracted. Of these, 97 studies were excluded for following reasons:

- studies based on unpublished data only = 3;
- duplicate studies = 40;
- studies presenting indirect evidence = 54.

This resulted in a final sample of 286 studies. Because a study can have multiple references, the 286 studies corresponded to three different sets of references (depicted in Figure 2):

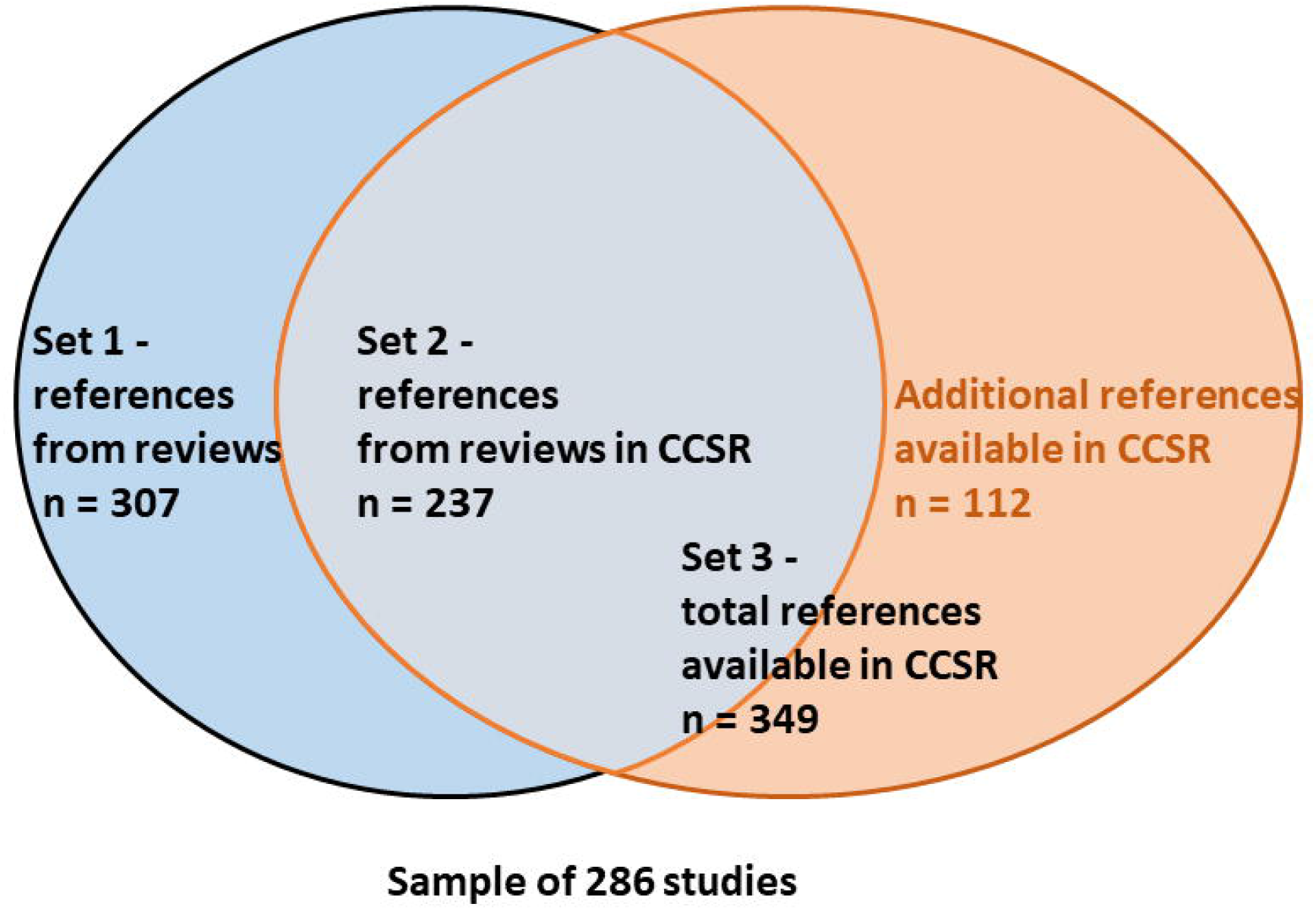

- a first set, consisting of 307 cited references from the reviews in the sample;
- a second set, consisting of 237 references available in CCSR from these 307 references;
- a third set, consisting of 349 references, which are the 237 references available in CCSR plus 112 additional references to the studies, which were only available in the CCSR, but not cited in the reviews.

As part of the evaluation, we also recorded how many of the 112 additional references in the CCSR would have been potentially ‘findable’ for review authors, if they had searched the CCSR. We recorded an additional reference as ‘findable’ if the date the reference was added to the CCSR preceded the search date of the review that cited the study.

In addition to calculating performance indicators, we conducted an audit trail of references not retrieved by the CCSR, incorrectly classified, incorrectly linked, or delayed in processing by > 30 days from the date they were added to the CRS segment.

## 3. Results

The original searches for the reviews in the sample were conducted between 4 April 2020 and 21 September 2020. 74% (212/286) of studies from the sample were retrieved by searches conducted before 1 July 2020. As the majority of studies on which our evaluation is based upon are from the first half of 2020, and no studies from after 21 September 2020, this report primarily reflects the performance of the CCSR for references published in the first half of 2020.

### 3.1. Sensitivity evaluation

The evaluation showed an overall sensitivity of 77.2% (237/307) for the CCSR. We present sensitivity results by CEOsys topic areas, publication type and study type in Table 2.

**Table 2:**
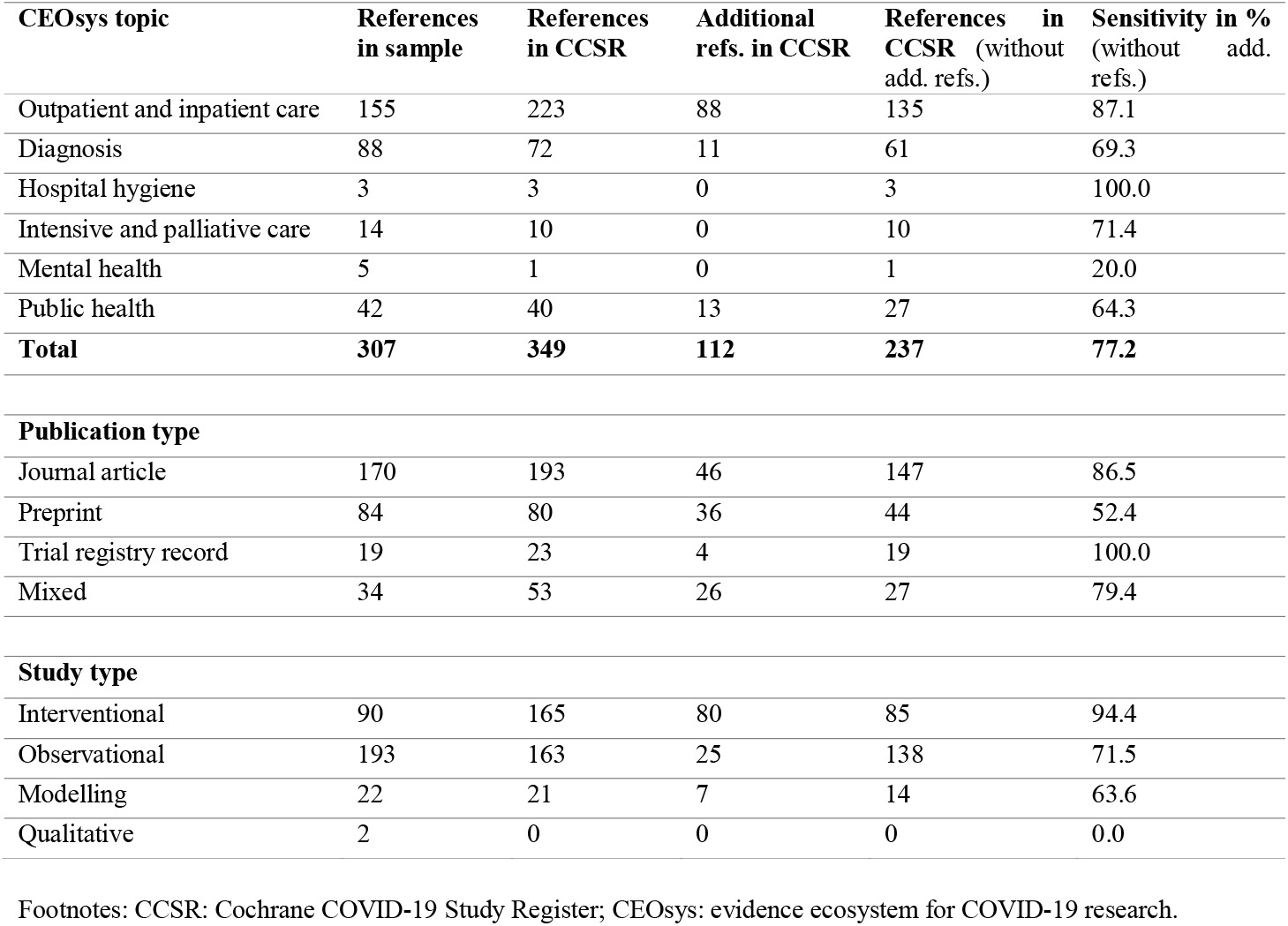
Sensitivity of CCSR per CEOsys topic, publication type, and study type.

| CEOsys topic | References in sample | References in CCSR | Additional refs. in CCSR | References in CCSR (without add. refs.) | Sensitivity in % (without add. refs.) |
| --- | --- | --- | --- | --- | --- |
| Outpatient and inpatient care | 155 | 223 | 88 | 135 | 87.1 |
| Diagnosis | 88 | 72 | 11 | 61 | 69.3 |
| Hospital hygiene | 3 | 3 | 0 | 3 | 100.0 |
| Intensive and palliative care | 14 | 10 | 0 | 10 | 71.4 |
| Mental health | 5 | 1 | 0 | 1 | 20.0 |
| Public health | 42 | 40 | 13 | 27 | 64.3 |
| **Total** | **307** | **349** | **112** | **237** | **77.2** |
| **Publication type** |  |  |  |  |  |
| Journal article | 170 | 193 | 46 | 147 | 86.5 |
| Preprint | 84 | 80 | 36 | 44 | 52.4 |
| Trial registry record | 19 | 23 | 4 | 19 | 100.0 |
| Mixed | 34 | 53 | 26 | 27 | 79.4 |
| **Study type** |  |  |  |  |  |
| Interventional | 90 | 165 | 80 | 85 | 94.4 |
| Observational | 193 | 163 | 25 | 138 | 71.5 |
| Modelling | 22 | 21 | 7 | 14 | 63.6 |
| Qualitative | 2 | 0 | 0 | 0 | 0.0 |

Footnotes: CCSR: Cochrane COVID-19 Study Register; CEOsys: evidence ecosystem for COVID-19 research.

The CCSR had the highest sensitivity (87.1%, 135/155) for the topic area of outpatient and inpatient care and the lowest for public health (64.3%, 27/42). As the number of references in the sample for the areas of hospital hygiene (n=3) and mental health (n=5) was very small, we considered these calculations unlikely to represent the true sensitivity of the CCSR in these areas.

The register had very high sensitivity for trial registry records (100%, 19/19) and high sensitivity for journal articles (86.5%, 147/170). The lowest sensitivity was for preprints (52.4%, 44/84). For the purposes of our analysis, we classified cited studies with multiple references of different publication types as being “mixed”. The CCSR had 79.4% (27/34) sensitivity for references with mixed publication types.

With regard to study type, we found the CCSR to have the best sensitivity for interventional study types (94.4%, 85/90) and the lowest for modelling studies (63.6%, 14/22). The number of qualitative studies in the sample (n=2) was not large enough to yield a representative calculation.

To better understand why references had not been captured, we conducted a sensitivity audit of all references, irrespective of topic area or study type, that were not retrieved by the CCSR (n=70). The audit data are publicly available.^7^ We found that 81.4% (57/70) had been retrieved by the search and added to the CRS segment, but not fully processed and published to CCSR, and that 18.6% (13/70) had not been retrieved by the centralised search processes. 78.5% (55/70) of the missed studies were observational and 11.4% (8/70) were modelling studies. Of the four interventional studies that were missed by the CCSR, all were preprints with other available references for the same studies in the CCSR. Two additional qualitative missed references were a preprint and a journal article for the same study.

Of the 57 references that were added to the CRS segment but not yet published to the CCSR, 57.8% (33/57) were preprints awaiting processing, 21% (12/57) were letters awaiting processing, 17.5% (10/57) were marked as not eligible and one as ‘unclear.’ One missing journal article had been captured by a repeated PubMed search and was awaiting processing.

Of the 13 references that were not added to the CRS segment, 76.9% (10/13) were preprints and 23.1% (3/13) were journal articles. Four of the preprints were the only available version of the study. Six were from medRxiv, but not captured by the centralised search process. Three were from SSRN and one was from preprints.org (both sources not currently searched for the register’
ss production). All three journal article references not added to the segment were the only available versions of the studies. Two were from an unclear source (we could not locate) and one was available in PubMed in April 2020 but missed by the search due to a known processing error.

### 3.2. Accuracy evaluation

The evaluation of the accuracy of classifications showed that 98.3% of the references were correctly classified with regard to study type (categories: interventional, observational, modelling, qualitative, other). With regard to study aim (categories: diagnostic/prognostic, treatment and management, health services research, epidemiology, transmission, mechanism, prevention, other) a total of 93.4% of references was accurately classified (Table 3).

**Table 3:** Accuracy of classifications in CCSR with regard to study type and study aim.

| CEOsys topic | References in CCSR | Correct study type | % | Correct study aim | % |
| --- | --- | --- | --- | --- | --- |
| Outpatient and inpatient care | 223 | 220 | 98.7 | 219 | 98.2 |
| Diagnosis | 72 | 69 | 95.8 | 69 | 95.8 |
| Hospital hygiene | 3 | 3 | 100.0 | 3 | 100.0 |
| Intensive and palliative care | 10 | 10 | 100.0 | 9 | 90.0 |
| Mental health | 1 | 1 | 100.0 | 0 | 0.0 |
| Public health | 40 | 40 | 100.0 | 26 | 65.0 |
| **Total** | **349** | **343** | **98.3** | **326** | **93.4** |

Footnotes: CCSR: Cochrane COVID-19 Study Register; CEOsys: evidence ecosystem for COVID-19 research.

The audit of misclassifications showed that six references were classified incorrectly with regard to study type as “observational” instead of “interventional” (audit data publicly available).^7^ Twenty-three references were incorrectly classified with regard to study aim. Thirteen should have been classified as “diagnostic/prognostic”, four as “transmission”, four as “treatment and management” and two as “modelling”.

With regard to the accuracy of study linking, 76 studies from the sample had more than one linked reference in the CCSR. In total, the CCSR contained 203 references for these 76 studies. 8 of 76 studies were found to have a linking error (11%). Thus, we found 89% of studies in the CCSR to be accurately linked.

When doing the analysis we became aware of an error in the proposed calculation of linking accuracy in our protocol. The denominator originally mentioned “studies with linked references in the sample”, but we intended to use studies with linked references in CCSR. This was corrected within this manuscript and analysed as such.

### 3.3. Currency evaluation

The publishing time is the time in days from the reference first becoming available in the original source until published in the CCSR. From 237 references analysed, 193 references (81.4%) were published to the register in under 30 days and 44 references (18.6%) were published to the register in over 30 days.

An assessment per publication type showed that 94.2% journal articles, 100% trial registry records were published in less than 30 days after being added to the original source, while 66% of preprints had a publishing time of over 30 days (Table 4).

**Table 4:** Publishing time of references to the CCSR.

| Publication type | References in sample available in CCSR | References published in < 30 days | % | References published in > 30 days | % |
| --- | --- | --- | --- | --- | --- |
| Journal article | 156 | 147 | 94.2 | 9 | 5.8 |
| Preprint | 53 | 18 | 34.0 | 35 | 66.0 |
| Trial registry record | 28 | 28 | 100.0 | 0 | 0.0 |
| **Total** | **237** | **193** | **81.4** | **44** | **18.6** |

Footnotes: CCSR: Cochrane COVID-19 Study Register.

An assessment of currency per publication type showed that the “time to segment” was quick (median of 4 days), with trial registry records taking one day, journal articles 3 days and preprints 5 days to being added to the segment. The “time from segment to register” was also quick for journal articles (median of 1 day) and trial registry records (median of 0 days), but preprints took over a month (median of 33 days) to be fully processed. Overall, the full publishing time was quick (median of 3 days), with 0.5 day for trial registry records and 2 days for journal articles, while taking around two months (median of 56 days) for preprints (Table 5).

**Table 5:** Time to add references to segment, to CCSR and total publishing time in days.

|  | Time to segment |  | Time from segment to register |  | Publishing time |  |
| --- | --- | --- | --- | --- | --- | --- |
|  | mean | median | mean | median | mean | median |
| Journal article | 4.5 | 3 | 6.5 | 1 | 11 | 2 |
| Preprint | 11 | 5 | 48.5 | 33 | 60 | 56 |
| Trial registry record | 1 | 1 | 1 | 0 | 1 | 0.5 |
| **Total** | **5.5** | **4** | **15** | **1** | **21** | **3** |

Footnotes: CCSR: Cochrane COVID-19 Study Register.

We audited 44 references that took over 30 days to publish (audit data publicly available).^7^ Thirty-five (79.5%) were preprints added after the May revision of the CCSR’s eligibility criteria to include this publication type. Of the 9 (20,5%) remaining journal article references, 7 were letters and 2 were full articles that were missed due to a processing error in April. All were captured by additional searches for letters and April publications identified as missing by the May evaluation.

### 3.4 Additional references

For each study from the sample, we recorded 112 additional references available in the CCSR. For each of the additional references, we compared the dates the references were added to the CCSR and the dates of search from the reviews. We found that 70 additional references out of the 112 were ‘findable’ in the CCSR by the dates of original searches of the reviews (see additional data publicly available).^7^

## 4. Discussion

The development work on the CCSR began in early March 2020. At this time, the volume of COVID-19 research was increasing dramatically. Cochrane saw the need for a study-based register to support rapid and living evidence synthesis that met Cochrane’s quality standards for review production. Therefore, the CCSR’s earliest search protocol reflected mandatory search sources in the Cochrane Handbook for Systematic Reviews of Interventions and the Methodological Expectations of Cochrane Intervention Reviews (MECIR).^8,9^ Hence, the Centralised Search Team focused their efforts on capturing COVID-19 studies from trial registers and bibliographic databases.

During the same period (in the early months of 2020), there was widespread publication of non-peer-reviewed preprints to online repositories and archives, such as medRxiv, bioRxiv, arXiv, Research Square, SSRN, RePEc, and others.^10,11^ An internal evaluation of the CCSR in mid-May 2020 revealed that preprints comprised nearly half of the included studies (184/373) from a sample of six COVID-19 reviews. The internal evaluation also found that 90% of the preprints included in the reviews were retrieved from medRxiv. To ensure the value of the CCSR, medRxiv was included as a source in May 2020.

Part of the CCSR’s production has been a learning process about supporting evidence synthesis on emerging infectious diseases with pandemic scope. Over the course of 2020, the Centralised Search Team witnessed the emergence of an evidence base that relied heavily on preprints and letters. We now recognise the importance of sources that are faster to publish and disseminate than full, peer-reviewed journal articles. Developers of future study reference collections or study-based registers on emerging infectious diseases should prespecify inclusion of non-traditional evidence sources, like preprints, and plan how to integrate them into information retrieval workflows.

While this evaluation shows that the CCSR has processing work to complete with regards to preprints and letters, 77.2% overall sensitivity is a significant improvement from the 39% sensitivity calculated in the early internal evaluation, particularly for the topic area of “outpatient and inpatient care” which shows a high sensitivity of 87.1%.

Granted that in May 2020 there were few interventional study references included in the analysis (n=20), the sensitivity at that time was only 60%. This current evaluation provided a larger sample (n=90) of interventional study references and found the sensitivity to be markedly higher at 94.4%. In addition, we saw that very few journal articles were missed by the CCSR and no trial registry records missed at all. Especially the latter finding is encouraging, because once a trial register record has been identified, the existence of a study is known, regardless of its publication status. Altogether this evaluation suggests the recent processes for capturing these publication types are working well.

In our sensitivity calculations we did not include the additional references contained in the CCSR for the sample studies that were not cited by the reviews in our sample. Nevertheless, the evaluation found 70 additional references which were available in the CCSR when original searches for the reviews in the sample were conducted. As potentially valuable data sources that could have impacted review findings, these available additional references illustrate the value of the CCSR’s study-based design. The CCSR’s public search interface captures all linked references for the study records retrieved, saving author time to identify and associate references for the same studies included in their reviews.

### 4.1 Limitations

This evaluation aimed to establish a representative sample set based on 20 reviews. This aim was achieved with regard to review type (five rapid, six living, and nine systematic reviews) and three of the six CEOsys topic areas (nine reviews on “outpatient and inpatient care”, four reviews on “diagnosis”, three reviews on “intensive and palliative care”). For the other three CEOsys topic areas, the sample selection only identified two reviews on “public health” and one each for “hospital hygiene” and “mental health”. This limitation is due to the low publication output in these fields during the period of the sample selection (July to November 2020). Because the validity for these three topic areas was limited, this evaluation can only draw conclusions on topic areas where the sample was deemed to be representative.

For the purposes of evaluating the currency of the CCSR’s production processes, a manual adjustment was made to the “date available” values for references published before the CCSR was launched on 1 April 2020. Any publication dates for references that predated the launch-date of the CCSR were adjusted to 1 April 2020. This ensured our currency measurements reflected true processing times and were not inflated to account for months of processing that could not have occurred prior to the CCSR’s inception.

Owing to the variability in searching methods in the reviews constituting the sample, our resource limitations and our aim to compare the results with the internal May 2020 evaluation, we did not replicate searches or rescreen results based on the sample reviews. It is a further limitation of the evaluation that we assumed all searches that could have been conducted in the CCSR would have been sufficiently sensitive to identify relevant studies available within the register. Future assessments of the CCSR’s public search interface are needed to determine user experience and to improve the likelihood of all audiences retrieving the maximum number of relevant references for their search queries.

## 5. Conclusion

This evaluation of the Cochrane Covid-19 Study Register (CCSR) was based on a sample of 20 COVID-19 reviews (rapid, living and systematic) published between 1 July and 12 November 2020. The sample yielded a set of 286 studies, which were used to evaluate the sensitivity, accuracy and currency of the study-based register.

Overall, the CCSR is performing well in all aspects. Particularly noteworthy are the high sensitivity of journal articles (86.5%) and trial registry records (100%), the accuracy of the study classifications (93.4-98.3%) and the study linking (89%). Preprints, which had already been identified as not being captured in a timely and sensitive manner in the early internal evaluation of the register, are now being identified in most cases, but have a markedly longer processing time in comparison to other publication types.

The CCSR provides access to additional references, which were not cited in the reviews forming the sample. Therefore, review producers can make additional use of the CCSR by checking their included studies in the register before publication of their reviews, in order to identify additional references added to the register after the initial search or not identified by the initial search in other databases.

Based on the high accuracy of the study classifications, the register can be considered especially useful for the production of rapid reviews. Researchers working on this review type can select relevant studies by limiting to study type and study aim, especially in the topic areas of “outpatient and inpatient care”, “diagnosis” and “intensive and palliative care”. For the process of evidence synthesis production within CEOsys, the CCSR has proven to be a time-saving resource providing a rapid overview of the available evidence base. Future research is needed to empirically estimate time savings to review producers through the use of information specialist maintained study-based registers.

## Data Availability

The data that support the findings of this study are available on OSF: https://osf.io/t3ycx/.

## Acknowledgements

This work was conducted within the CEOsys project (www.covid-evidenz.de), funded under a scheme issued by the Network of University Medicine (Nationales Forschungsnetzwerk der Universitätsmedizin (NUM)) by the Federal Ministry of Education and Research of Germany (Bundesministerium für Bildung und Forschung (BMBF)), grant number 01KX2021.

We would like to acknowledge Jaqueline Hildebrandt for her help with parts of the data extraction and Isabelle Boutron, Chris Mavergames, Steve McDonald, Jörg Meerpohl, Anna Noel-Storr, Bernd Richter and Susi Wisniewski for their comments.

## Data availability statement

The data that support the findings of this study are available on OSF: https://osf.io/t3ycx/.

## Highlights

- What is already known
  - In early 2020, there was a publishing surge of COVID-19 studies. High publication rates of trial registry records, preprints and journal articles on COVID-19 studies persisted through the year.
  - Well maintained study-based registers facilitate evidence synthesis production.
  - The Cochrane COVID-19 Study Register (CCSR) was launched on 1 April 2020 as a freely-available, continually-updated, annotated reference collection of human primary studies on COVID-19 with an aim to support rapid evidence synthesis.
- What is new
  - The evaluation of the CCSR shows that it is perfoming well in collecting and continually updating the evidence base on COVID-19 with regard to sensitivity, accuracy and currency.
  - Efforts to maintain a study-based register are worthwhile for rapid and living evidence synthesis production and can assist in the identification of additional references for included studies.
  - There is an important role for preprints in evidence synthesis on emerging infectious diseases. COVID-19 related reviews produced during the first half of 2020 relied heavily on preprints.
- Potential impact for *Research Synthesis Methods* readers outside the authors’ field
  - Information specialists play a role in evidence synthesis production by developing and maintaining study-based registers, which can be used to streamline several evidence synthesis endeavours.

