## Supplementary file 1 for "Evaluation of the sensitivity, accuracy and currency of the Cochrane COVID-19 Study Register for supporting rapid evidence synthesis production"

### Supplementary file 1: Appendix

**PubMed search strategy to identify reviews for the sample, run on 21 November 2020.**

(2019 nCoV[tiab] OR 2019nCoV[tiab] OR corona virus[tiab] OR corona viruses[tiab] OR coronavirus[tiab] OR coronaviruses[tiab] OR COVID[tiab] OR COVID19[tiab] OR nCov 2019[tiab] OR SARS-CoV2[tiab] OR SARS CoV-2[tiab] OR SARSCoV2[tiab] OR SARSCoV-2[tiab] OR "Coronavirus"[Mesh:NoExp] OR "COVID-19"[nm] OR "COVID-19 drug treatment"[nm] OR "COVID-19 diagnostic testing"[nm] OR "COVID-19 serotherapy"[nm] OR "COVID-19 vaccine"[nm] OR "LAMP assay"[nm] OR "severe acute respiratory syndrome coronavirus 2"[nm] OR "spike protein, SARS-CoV-2"[nm]) NOT ("animals"[mh] NOT "humans"[mh]) NOT (editorial[pt] OR newspaper article[pt]) AND (meta-analysis[Filter] OR systematicreview[Filter]) AND ("Nat Rev Dis Primers"[Journal] OR "JAMA Intern Med"[Journal] OR "J Cachexia Sarcopenia Muscle"[Journal] OR "PLoS Med"[Journal] OR "J Travel Med"[Journal] OR "Cochrane Database Syst Rev"[Journal] OR "BMJ"[Journal] OR "CMAJ"[Journal] OR "JAMA"[Journal] OR "N Engl J Med"[Journal] OR "Ann Intern Med"[Journal] OR "Lancet"[Journal] OR "JAMA Netw Open"[Journal] OR "Med J Aust"[Journal] OR "Am J Med"[Journal] OR "Mayo Clin Proc"[Journal] OR "J R Soc Med"[Journal] OR "Am J Prev Med"[Journal] OR "J Gen Intern Med"[Journal] OR "J Intern Med"[Journal] OR "Eur J Intern Med"[Journal] OR "Br J Gen Pract"[Journal] OR "Amyloid"[Journal] OR "Ann Fam Med"[Journal] OR "BMC Med"[Journal] OR "Transl Res"[Journal] OR "Dtsch Arztebl Int"[Journal]) AND ("2020/07/01"[dp] : "3000"[dp])
